# The ADEPT Study, A Comparative Study of Dentists’ Ability to Detect Enamel-only Proximal Caries in Bitewing Radiographs With and Without the use of AssistDent^®^ Artificial Intelligence Software

**DOI:** 10.1101/2020.10.12.20211292

**Authors:** Hugh Devlin, Tomos Williams, Jim Graham, Martin Ashley

## Abstract

**Introduction:** Reversal of enamel-only proximal caries by non-invasive treatments is important in preventive dentistry. However, detecting such caries using bitewing radiography is difficult, and the subtle patterns are often missed by dental practitioners.

**Aims:** To investigate whether the ability of dentists to detect enamel-only proximal caries is enhanced by the use of AssistDent^®^ Artificial Intelligence (AI) software.

**Materials and Methods:** In the ADEPT (**<u>A</u>**ssist**<u>D</u>**ent **<u>E</u>**namel-only **<u>P</u>**roximal caries assessmen**<u>T</u>**) study, twenty-three dentists were randomly divided into a control arm, without AI assistance, and an experimental arm in which AI assistance provided on-screen prompts indicating potential enamel-only proximal caries. All participants analysed a set of 24 bitewings in which an expert panel had previously identified 65 enamel-only carious lesions and 241 healthy proximal surfaces.

**Results:** The control group found 44.3% of the caries, whereas the experimental group found 75.8%. The experimental group incorrectly identified caries in 14.6% of the healthy surfaces compared to 3.7% in the control group. The increase in sensitivity of 71% and decrease in specificity of 11% are statistically significant (p<0.01).

**Conclusions:** AssistDent^®^ Artificial Intelligence software significantly improves dentists’ ability to detect enamel-only proximal caries and could be considered as a tool to support preventive dentistry in general practice.

**Key Points:** Enamel-only proximal caries are often missed by dentists when examining bitewing radiographs.

The use of AssistDent^®^ Artificial Intelligence software results in a 71% increase in ability to detect enamel-only proximal caries accompanied by a 11% decrease in specificity.

Artificial Intelligence software could be considered as a tool to support preventive dentistry in general practice.

## Introduction

The early detection and treatment of enamel-only proximal caries can preserve tooth structure and prevent the subsequent cycle of treatment and re-treatment that is involved with more invasive treatment. Recent guidelines from the NHS encourage preventive care in dental practices, especially for young children. Patients differ widely on their willingness to pay for preventive therapies^1^ but nearly all parents value a healthy dentition for their children and are willing to invest resources to maintain this.^2^ However, it is well documented that preventive care in children and adults in the UK is offered less frequently than it should be and therefore if prevention is to be adopted more widely in adults, caries detection must be time efficient and accurate. Only then can the ideal, personalised caries assessment of adult patients and their preventive care be developed.

Radiographic examinations can increase the number of carious lesions that are detected over those that would be detectable by clinical examination alone. The Department of Health recommends that dentists use the FGDP (UK) guideline document ‘*Selection Criteria for Dental Radiography*’ in determining the frequency of use of bitewing radiography.^3^ Nevertheless, systematic reviews have consistently shown that detection of proximal caries on bitewing radiography has a low sensitivity.^4^ A number of studies have reported poor diagnostic sensitivity for radiographic detection of demineralisation by dentists. In a classic study by Mejàre et al.^5^, premolar and adjacent teeth surfaces were examined radiographically and visually. The premolar teeth were then extracted for orthodontic reasons. They found that the sensitivity of detection of enamel-only proximal caries was 36.7%. Other studies have found similarly low sensitivity values.^6,7^

The introduction of Artificial Intelligence (AI) methods allows routine tasks to be conducted more quickly and efficiently. AssistDent^®^ is an AI software product, developed by Manchester Imaging Limited, and uses machine learning algorithms to search for evidence of enamel-only proximal caries on bitewing radiographs.^8^ It is an aid to the dentist, assisting their clinical decision-making by providing on-screen prompts for potential locations of enamel-only proximal caries. The final judgement about whether enamel-only proximal caries is present, or not, is a decision for the clinician.

The null hypothesis of this research was that there is no difference in the performance of dentists in diagnosing the presence of enamel-only proximal caries on bitewing radiographs with and without the use of AssistDent^®^.

## Methods

A pilot study ^9^ was conducted with dental students to assist in developing the methodology and provide initial data for a sample size calculation. The final study protocol, participant information sheet and consent forms for the study were approved by the Manchester University Research Ethics Committee (Ref: 2020-9892-15955).

Participants were recruited from two sources: 1) dentists practising as general dental practitioners who in addition provide tutorage for dental students within the University of Manchester Dental School; 2) practising dentists undertaking postgraduate training within the University of Manchester Foundation NHS trust. All signed informed consent forms.

The dentists were randomly divided into control and experimental arms by pairing the participants within the recruitment sources according to the order in which they were enrolled. The first of each pair of participants were randomly assigned to either the control or experimental arm with the second assigned to the other group. This method ensured random assignment while maintaining even arm sizes equally balanced between the recruitment sources as the study progressed.

Both arms examined the same images using the same graphical user interface as shown in figure 1. In the control group (n=11), the caries prompting function of AssistDent^®^ was disabled in order to measure the ability of the group to detect enamel-only proximal caries without the use of AI software. In the experimental group (n=12), the caries prompting function of AssistDent^®^ was enabled in order to assist the participants.

**Figure 1.**
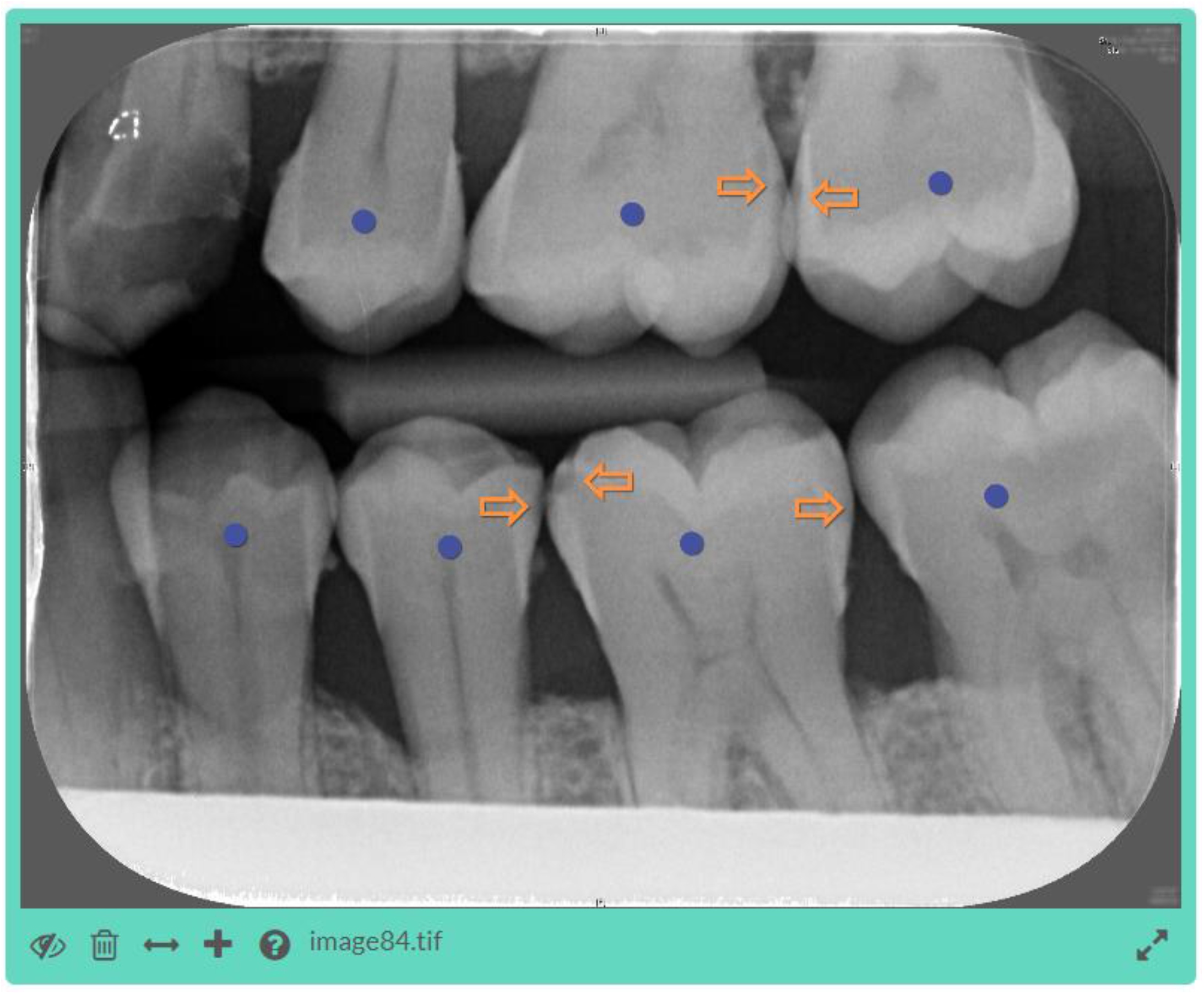
The AssistDent^®^ Graphical User Interface. Orange arrows indicate the presence of enamel-only proximal caries and purple circles indicate that the tooth has been detected and analysed by the AI algorithm. The experimental group were provided with an initial set of indicators generated by the AI software while no initial indicators were generated for the control group. Controls in the lower left menu enable users to add and delete indicators. Indicators can be moved by dragging them to the desired position. Once the analysis of the image is complete and the user is satisfied that every enamel-only proximal caries is marked by an arrow, the participant pressed the save button, bottom left, and proceeded to the next image.

So that the study would have applicability to general practice, a total of 1,446 bitewing radiographs were collected from a range of different sites (1 teaching hospital site and 9 general dental practitioner (GDP) sites). Separate ethical approval had been received from the Integrated Research Application System (IRAS project ID: 248306, REC reference: 18/NI/0111). As illustrated in figure 2, a test set of 103 images were selected randomly, but with the same proportion across the image acquisition sites, and excluded from all AI model training and evaluation. A further subset of 24 images were selected form the test set for the study, again representative of the acquisition sites but with the criterion that there was at least one enamel-only proximal caries in each image. Images from one of the GDP sites were excluded due to their poor quality. Another GDP site lacked images with one enamel-only proximal caries, therefore two study images from this site had no enamel-only proximal caries. For practical purposes, the prevalence of caries was higher in our study set than the general population. However, this did not affect the measures of sensitivity and specificity. The images were presented to each participant in the same order, grouped according to the acquisition site.

**Figure 2.**
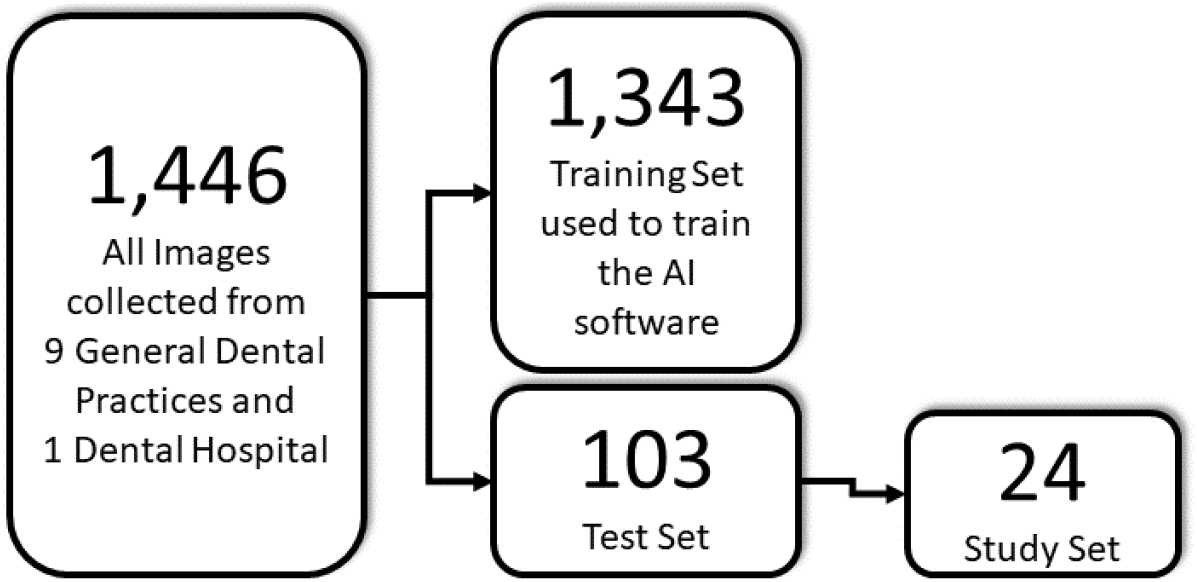
Selection of images for the study. All images were divided into a training and test set, with the test set excluded from the training and evaluation of the AI models. A subset of training set images was chosen for the study.

Gold Standard annotation of all classes of proximal caries was obtained from a panel of five dento-maxillofacial radiologists and one professor of Restorative Dentistry, each of whom annotated the location and grade of caries on a set of images independently of each other. Each image was annotated by at least three members of the panel. These individual expert annotations were consolidated by retaining in the gold standard set any region identified by any expert as enamel-only proximal caries while removing duplicate annotations, resulting in a gold standard set of 1,972 examples of enamel-only proximal caries for algorithm training and evaluation.

The caries annotations entered by each participant were collected remotely via a web application and analysed to determine whether they were true positives (correct identifications of enamel-only proximal caries) or false positives (annotations not corresponding to the location of the gold standard enamel-only proximal caries). Annotations corresponding to dentine proximal caries were recorded but excluded from this analysis. The **True Positive Rate** or **Sensitivity** of diagnosis is a measure of how well a participant detected the enamel-only proximal caries and was calculated as the sum of true positives divided by the sum of the gold standard caries. **True Negative Rate** or **Specificity** is a measure of how well the participant identified healthy surfaces and did not mark them as carious. The probability of a false detection is quoted in terms of a **False Positive Rate** calculated as the sum of false positive detections divided by the sum of healthy surfaces, which is equal to 1-Specificity.

## Results

A per-participant breakdown of the evaluation scores and performance measures, together with the aggregate scores and measures for each arm, is presented in the Supplementary Material. The data demonstrates that 23 dentists were recruited, 11 in the control arm and 12 in the experimental arm. These were balanced within the arms between the two recruitment sources of general dental practitioners and practising dentists undertaking postgraduate training. All participants analysed all 24 images.

Figure 3 presents the mean true positive and true negative rates over all participants and the 95% confidence intervals, for each arm. The improved mean true positive rate of the experimental arm participants (75.8% with AssistDent^®^) compared to the control arm (44.3% without AssistDent^®^) is clearly visible. This is accompanied by a decrease in true negative rate from 96.3% to 85.4%. Table 1 presents the statistical analysis for the performance measures within each arm together with the result of a student t-test. The t-tests demonstrate that the improved true positive rate and reduced true negative rate of the experimental compared to the control arm were significant with p-values below alpha of 0.01.

**Table 1.**
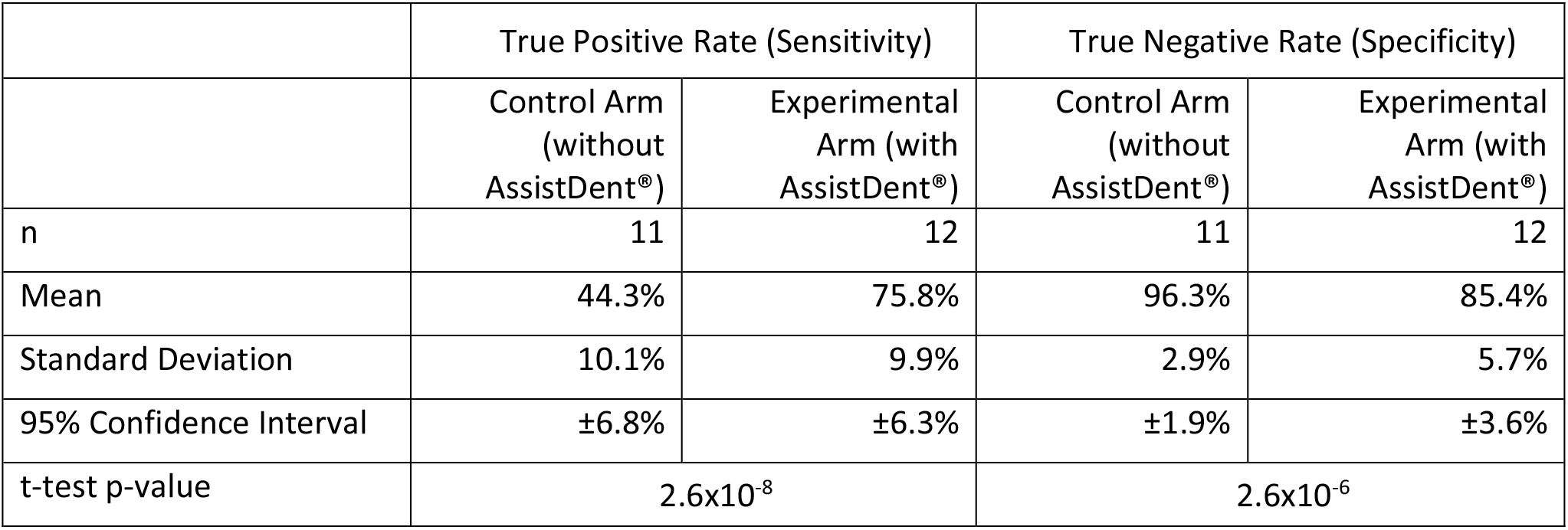
Statistical analysis of the mean per-participant performance measures for each arm.

|  | True Positive Rate (Sensitivity) |  | True Negative Rate (Specificity) |  |
| --- | --- | --- | --- | --- |
|  | Control Arm<br>(without<br>AssistDent®) | Experimental<br>Arm (with<br>AssistDent®) | Control Arm<br>(without<br>AssistDent®) | Experimental<br>Arm (with<br>AssistDent®) |
| n | 11 | 12 | 11 | 12 |
| Mean | 44.3% | 75.8% | 96.3% | 85.4% |
| Standard Deviation | 10.1% | 9.9% | 2.9% | 5.7% |
| 95% Confidence Interval | ±6.8% | ±6.3% | ±1.9% | ±3.6% |
| t-test p-value | 2.6x10<sup>-8</sup> |  | 2.6x10<sup>-6</sup> |  |

**Figure 3.**
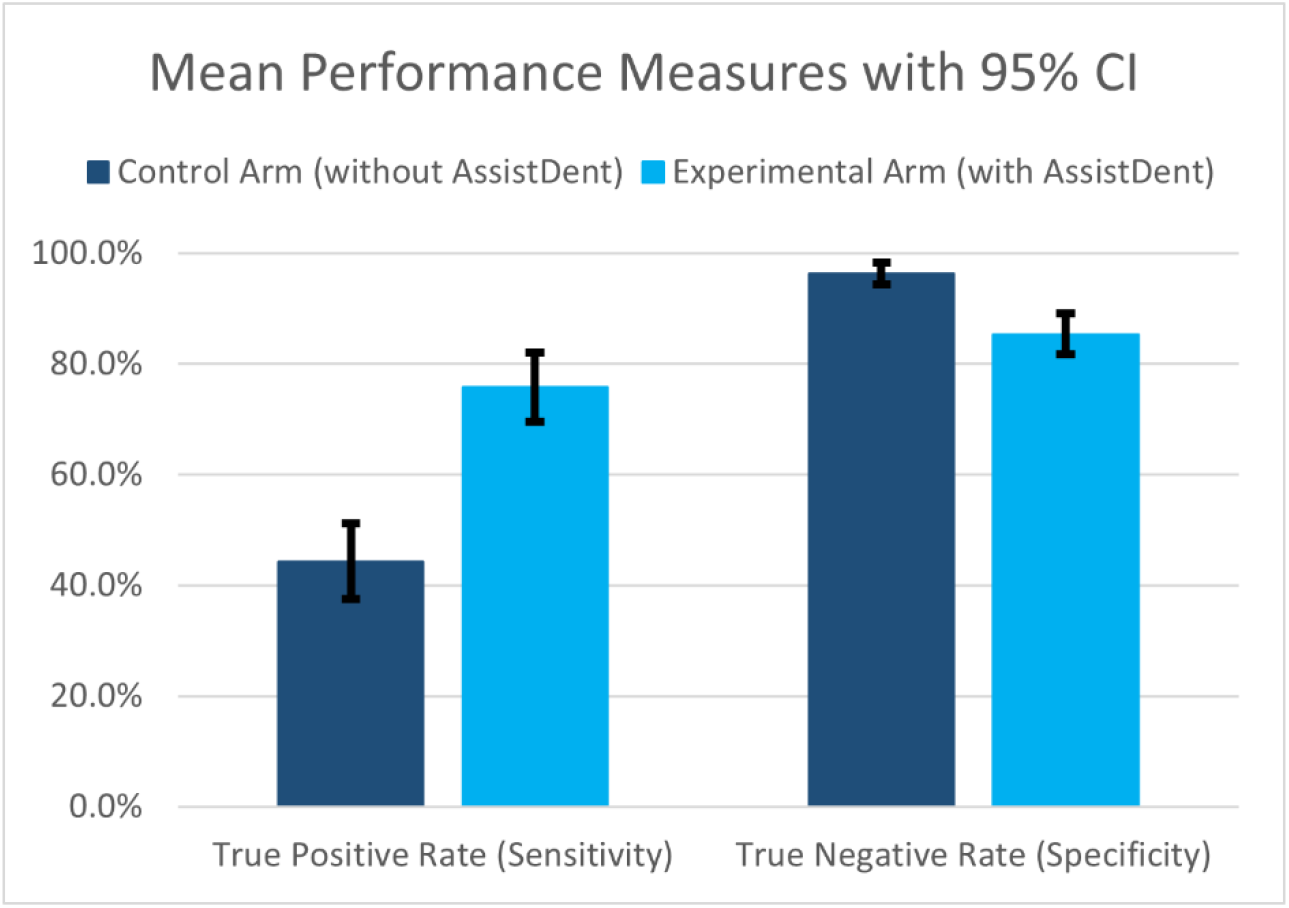
Bar charts showing the mean true positive and true negative rates together with their 95% confidence intervals, for each arm.

Table 2 presents the results of an odds-ratio comparison of true positive and true negative rates for each arm. The ratio between the experimental and control groups with a value greater than one indicates that use of AssistDent^®^ increased the ability to detect enamel-only proximal caries by 71%. Similarly, the ratio of less than 1 for true negative rate indicates that the experimental arm participants were 11% less likely to correctly identify healthy proximal surfaces as non-carious. A per-participant and per-group break-down of the results are presented in the supplementary material together with additional summary statistics for each group.

**Table 2.**
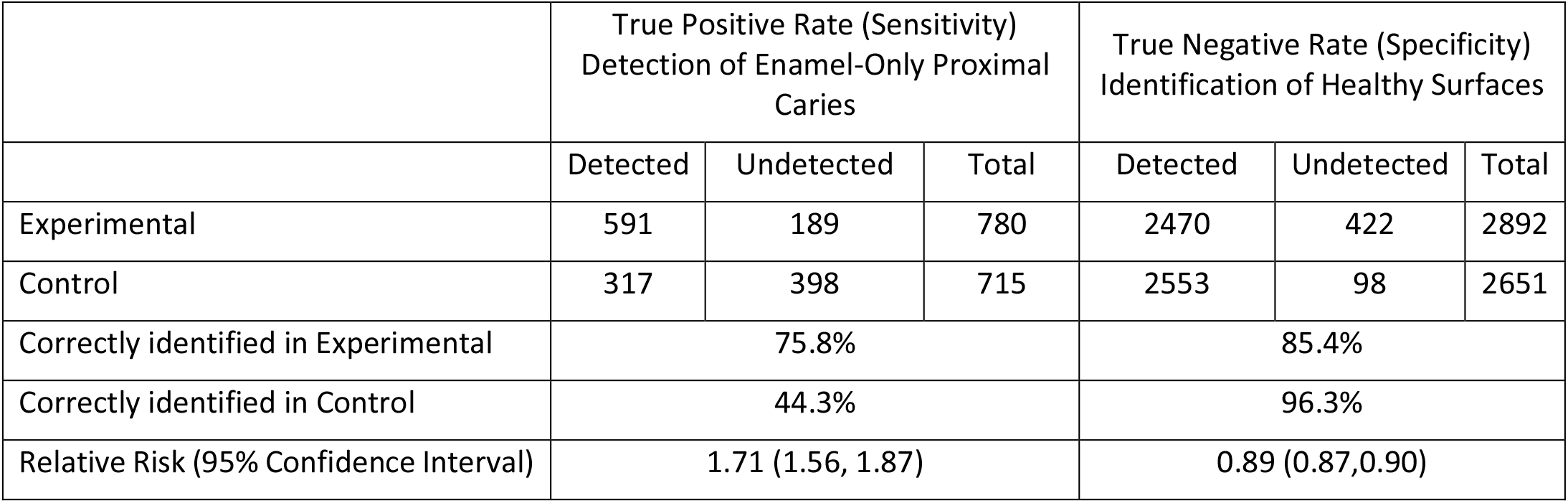
Odds-Ratio of true positive and true negative rates of the experimental group compared to the control group with 95% confidence intervals.

|  | True Positive Rate (Sensitivity)<br>Detection of Enamel-Only Proximal<br>Caries |  |  | True Negative Rate (Specificity)<br>Identification of Healthy Surfaces |  |  |
| --- | --- | --- | --- | --- | --- | --- |
|  | Detected | Undetected | Total | Detected | Undetected | Total |
| Experimental | 591 | 189 | 780 | 2470 | 422 | 2892 |
| Control | 317 | 398 | 715 | 2553 | 98 | 2651 |
| Correctly identified in Experimental | 75.8% |  |  | 85.4% |  |  |
| Correctly identified in Control | 44.3% |  |  | 96.3% |  |  |
| Relative Risk (95% Confidence Interval) | 1.71 (1.56, 1.87) |  |  | 0.89 (0.87,0.90) |  |  |

## Discussion

Small areas of proximal surface enamel demineralization are difficult to detect visually on radiographs.^10^ In a review of diagnostic studies reported by Keenan and Keenan,^11^ they found lower sensitivities for the detection of proximal lesions in clinical studies than in vitro studies. The mean sensitivity in the clinical studies was 0.24 and for the in vitro studies was 0.43.

Other studies that have measured sensitivity and specificity of caries detection using radiographs have found a large variation. This may be due to variation in the range of caries depth or the use of in vitro studies. In our in vivo study, we mainly used a sample of radiographs from a range of dental practices that were taken in the course of the examination of patients. The low sensitivity and high specificity of our control group in detecting caries on bitewing radiographs are comparable with the above studies, and with that reported in a systematic review.^12^

Low sensitivity in the detection of enamel-only caries is problematic in the context of preventive dentistry. A significant increase in sensitivity is required over the values reported in these studies, even at the expense of an increased number of false positive detections. Enamel lesions are treated in a conservative manner with dietary and interdental cleaning advice, fluoride treatments and potentially, resin infiltration. This, in conjunction with cooperation from the patient, will prevent operative intervention and the entry of the tooth into a restorative cycle of increasingly larger restorations and eventual extraction. An increase in sensitivity of detection is important as it provides a better assessment of the full extent of the patient’s caries experience. The dentist should create an individually tailored dental care plan with the patient based on a true picture of their oral health. Some aspects of this risk assessment are based on the findings from bitewing radiographs obtained during the process of screening for early, enamel-only caries. In those individuals who suddenly develop many early lesions, this would merit an investigative effort to determine any underlying medical or social causes followed up by more frequent recall intervals. The number of lesions present is an important variable when predicting future caries experience.^18^ However, increased sensitivity carries with it the likelihood of an increased number of false positive detections. This trade-off is inevitable, but it is clearly desirable that the false positive count should be kept as low as possible. The risk to patients arising from false positive detections is minimal provided clinicians follow guidelines and do not restore teeth with enamel-only caries. In a situation where a false positive detection is made when no caries is present, an individual may, as a result, receive preventive advice without any obvious clinical benefit. The decrease in specificity observed from the use of AssistDent accompanies a much larger increase in sensitivity.

There has been a growing interest in general dental publications and online fora in the use of AI in dentistry. A recent review by Schwedicke et al ^13^ has described the basics of AI and explored its potential use in diagnostics, treatment planning and conduct in dentistry. Computer image analysis is a particular application of AI in dentistry (and medicine more generally). An earlier, scoping review ^14^ refers particularly to the application in image diagnostics. There is a relatively small number of studies related to caries detection. For example, Lee et al.^15^ investigated the performance of a convolutional neural network on classification of caries in images of individual teeth isolated from periapical images. The study did not focus specifically on enamel-only caries. Srivastra et al.^16^ also trained a convolutional neural network for fully automatic detection of caries in bitewing images.

To our knowledge, AssistDent^®^ is the only commercially available AI system for use in the clinical diagnosis of enamel-only proximal caries, acting as a prompting system to support dentists’ diagnostic decisions. Schwendicke et al ^14^ recommend that “…the dental community should appraise [the AI systems] against the rules of evidence-based practice.” This study is an example of such an appraisal.

It is important that dentists receive appropriate training with any new diagnostic system. Qudeimat et al.^17^ investigated the effect of ICDAS training and found a significant increase in overtreatment recommendations. The reproducibility of any diagnostic system depends on the experience of the clinician and this is especially so where the diagnosis is a visual score rather than AI-based.^18^ How should our profession address the issue of poor sensitivity in detection of enamel-only proximal caries? Audit, reflection with peer review and evaluation of past performance are essential parts of dental practice. An audit of caries diagnosis will rely on identifying opportunities for improvement, comparison with an accepted standard of care and implementing change. The increased sensitivity arising from AI-supported detection may provide a useful standard for audit in caries detection. Testing will also reveal clinicians with unacceptable variation in bitewing analysis competency assessments, with direction towards further training. More efficient caries detection may help in identifying patients with higher caries risk, while the associated display of detected caries can provide a basis for encouraging a detailed discussion with the patient of their oral health and the factors affecting it. The intuitive graphical screen display of AssistDent^®^ may be more readily understood by patients in comparison to other algorithm-based tools such as the Cariogram. The latter uses a pie-diagram of important factors to illustrate the probability that future caries may be prevented.^19,20^ However, once the high caries risk individuals are identified, it is important to further investigate all the factors predisposing to caries such as a high sugar intake and infrequent brushing.^21^

Caries assessment by dentists using AssistDent^®^ is compatible with the ICCMS caries management system of the ICDAS Foundation and could be used in conjunction with it. AssistDent^®^ provides feedback on the proximal enamel surfaces using radiographs whereas ICCMS is a mainly visual assessment of the non-proximal surfaces for caries. Both systems aim to maintain tooth structure and encourage preventive care by developing a caries assessment for each patient. At this stage of its development, this AI software is used as an aid to diagnosis, but future developments could include monitoring the progression of caries.

## Conclusion

AssistDent^®^ Artificial Intelligence software increases dentists’ sensitivity when assessing enamel-only proximal caries. This increased sensitivity is accompanied by a smaller decrease in specificity. The increase in false positive diagnoses may occasionally result in unnecessary preventive treatment and associated use of limited healthcare resources.

The increase in sensitivity in detecting enamel-only proximal caries should enable better informed targeting of preventive treatments. This would contribute to avoiding the requirement for later restorative treatment, resulting in an overall saving of resources and an improvement in the dentition of patients.

## Data Availability

All data is provided in the main paper

## Declaration of interests

HD, JG and TW are employees, of Manchester Imaging Ltd. The Division of Dentistry, University of Manchester, purchased a software licence for AssistDent^®^ from Manchester Imaging Ltd. MA is not an employee of Manchester Imaging Ltd and declares no conflict of interest.

## Supplementary Material

### Per-participant breakdown of the evaluation scores and performance measures

Per-participant breakdown of the evaluation scores and performance measures together with the aggregate scores and measures for each arm. The aggregate measures presented at the bottom of the table are calculated as the sum of the quantitative measures across all participants within each arm, together with the aggregate performance measures. Every participant analysed all 24 images meaning that they were all exposed to 65 enamel-only proximal caries and 241 healthy surfaces.

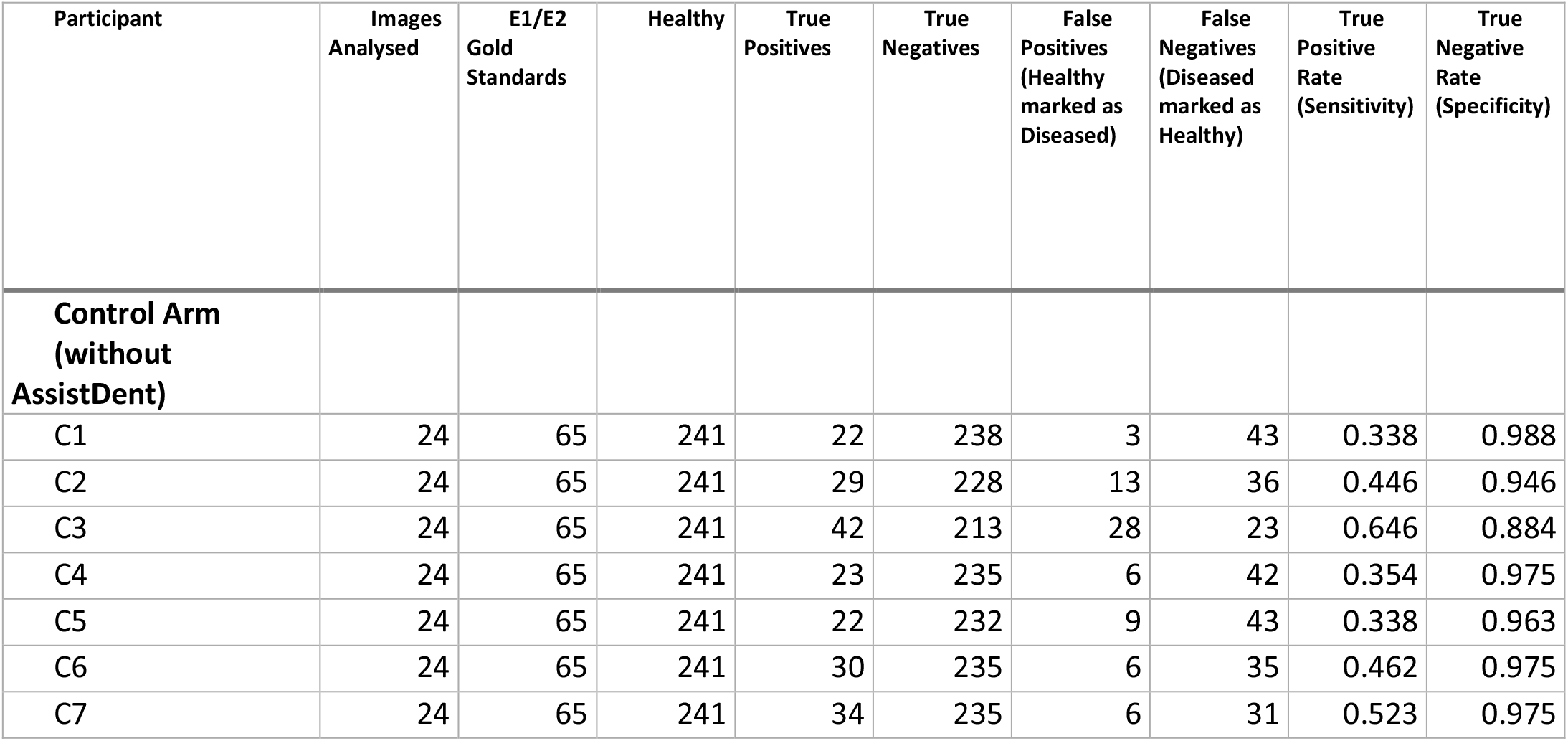
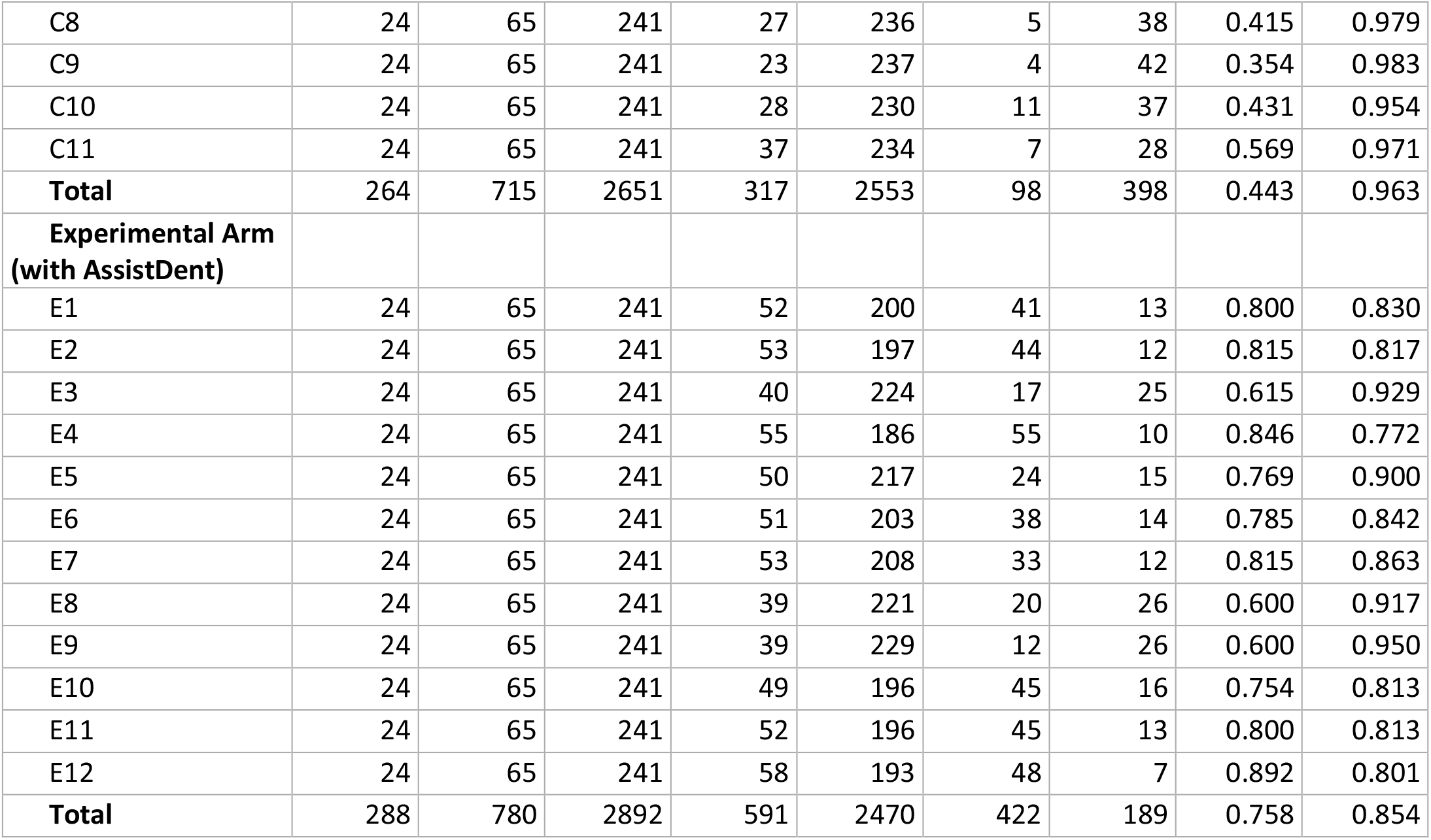

| Participant | Images Analysed | E1/E2 Gold Standards | Healthy | True Positives | True Negatives | False Positives (Healthy marked as Diseased) | False Negatives (Diseased marked as Healthy) | True Positive Rate (Sensitivity) | True Negative Rate (Specificity) |
| --- | --- | --- | --- | --- | --- | --- | --- | --- | --- |
| **Control Arm (without AssistDent)** |  |  |  |  |  |  |  |  |  |
| C1 | 24 | 65 | 241 | 22 | 238 | 3 | 43 | 0.338 | 0.988 |
| C2 | 24 | 65 | 241 | 29 | 228 | 13 | 36 | 0.446 | 0.946 |
| C3 | 24 | 65 | 241 | 42 | 213 | 28 | 23 | 0.646 | 0.884 |
| C4 | 24 | 65 | 241 | 23 | 235 | 6 | 42 | 0.354 | 0.975 |
| C5 | 24 | 65 | 241 | 22 | 232 | 9 | 43 | 0.338 | 0.963 |
| C6 | 24 | 65 | 241 | 30 | 235 | 6 | 35 | 0.462 | 0.975 |
| C7 | 24 | 65 | 241 | 34 | 235 | 6 | 31 | 0.523 | 0.975 |
| C8 | 24 | 65 | 241 | 27 | 236 | 5 | 38 | 0.415 | 0.979 |
| C9 | 24 | 65 | 241 | 23 | 237 | 4 | 42 | 0.354 | 0.983 |
| C10 | 24 | 65 | 241 | 28 | 230 | 11 | 37 | 0.431 | 0.954 |
| C11 | 24 | 65 | 241 | 37 | 234 | 7 | 28 | 0.569 | 0.971 |
| **Total** | **264** | **715** | **2651** | **317** | **2553** | **98** | **398** | **0.443** | **0.963** |
| **Experimental Arm**<br>**(with AssistDent)** |  |  |  |  |  |  |  |  |  |
| E1 | 24 | 65 | 241 | 52 | 200 | 41 | 13 | 0.800 | 0.830 |
| E2 | 24 | 65 | 241 | 53 | 197 | 44 | 12 | 0.815 | 0.817 |
| E3 | 24 | 65 | 241 | 40 | 224 | 17 | 25 | 0.615 | 0.929 |
| E4 | 24 | 65 | 241 | 55 | 186 | 55 | 10 | 0.846 | 0.772 |
| E5 | 24 | 65 | 241 | 50 | 217 | 24 | 15 | 0.769 | 0.900 |
| E6 | 24 | 65 | 241 | 51 | 203 | 38 | 14 | 0.785 | 0.842 |
| E7 | 24 | 65 | 241 | 53 | 208 | 33 | 12 | 0.815 | 0.863 |
| E8 | 24 | 65 | 241 | 39 | 221 | 20 | 26 | 0.600 | 0.917 |
| E9 | 24 | 65 | 241 | 39 | 229 | 12 | 26 | 0.600 | 0.950 |
| E10 | 24 | 65 | 241 | 49 | 196 | 45 | 16 | 0.754 | 0.813 |
| E11 | 24 | 65 | 241 | 52 | 196 | 45 | 13 | 0.800 | 0.813 |
| E12 | 24 | 65 | 241 | 58 | 193 | 48 | 7 | 0.892 | 0.801 |
| **Total** | **288** | **780** | **2892** | **591** | **2470** | **422** | **189** | **0.758** | **0.854** |

### Summary Statistics

Summary statistics for each group. The positive likelihood ratios for both groups are well above 1 while the negative likelihood ratio is below 1.

|  | **Control Arm (without**<br>**AssistDent)** | **Experimental Arm**<br>**(with AssistDent)** |
| --- | --- | --- |
| **Images Analysed** | 264 | 288 |
| **E1/E2 Gold Standards** | 715 | 780 |
| **Healthy** | 2651 | 2892 |
| **True Positives** | 317 | 591 |
| **True Negatives** | 2553 | 2470 |
| **False Positives (Healthy marked as Diseased)** | 98 | 422 |
| **False Negatives (Diseased marked as Healthy)** | 398 | 189 |
| **True Positive Rate (Sensitivity)** | 0.443 | 0.758 |
| **True Negative Rate (Specificity)** | 0.963 | 0.854 |
| **False Positive Rate (1-Specificity)** | 0.037 | 0.146 |
| **Positive Likelihood Ratio (Sensitivity / 1-Specificity)** | 11.993 | 5.193 |
| **Negative Likelihood Ratio (1 - Sensitivity / Specificity)** | 0.578 | 0.284 |
| **Accuracy** | 0.853 | 0.834 |
| **Positive Predictive Value** | 0.764 | 0.583 |
| **Negative Predictive Value** | 0.865 | 0.929 |

## Notes

### Clinical Trial

Manchester University Research Ethics Committee (Ref: 2020-9892-15955)

### Funding Statement

No external funding was received

### Author Declarations

Manchester University Research Ethics Committee

### Summary of Updates

Clarification throughout paper. Additional supplementary data.

